# A non-systematic signal-correction error in a commercial multiple-breath washout device significantly impacts outcomes in children and adults

**DOI:** 10.1101/2022.01.27.22269250

**Authors:** Marc-Alexander Oestreich, Florian Wyler, Blaise Etter, Kathryn A. Ramsey, Philipp Latzin

**Author notes:** **Corresponding author**: Philipp Latzin, MD, PhD, Inselspital, Bern University Hospital, Freiburgstrasse 15, CH-3010 Bern, Switzerland. Authors contributed equally to this work as first author (Marc-Alexander Oestreich, Florian Wyler).

## Abstract

**Background:** Multiple-breath nitrogen washout is an established and sensitive technique to assess functional residual capacity and ventilation inhomogeneity in the lung. Indirect calculation of nitrogen concentration requires precise calibration and accurate measurement of gas concentrations.

**Aim:** We investigated the accuracy of the carbon dioxide gas concentration used for the indirect calculation of nitrogen concentration in a commercial multiple-breath washout (MBW) device (EasyOne Pro LAB, ndd Medizintechnik AG, Zurich, Switzerland) and its impact on outcomes.

**Methods:** A high-precision calibration gas mixture was used to evaluate sensor output and calculated carbon dioxide concentration. We assessed the impact of a corrected CO_2_ signal on MBW outcomes in a dataset of healthy children and adults and children with lung disease.

**Results:** The EasyOne device uses a respiratory quotient-based adjustment to correct the measured carbon dioxide signal for potential long-term changes in sensor output. In the majority of measurements (89%), this resulted in an overestimation of expired nitrogen concentrations (range -0.7 to 1.7%), and consequently MBW outcomes. Correction of the CO_2_ signal reduced the mean (range) cumulative expired volume by 27.1% (−58.1%; 1.0%), functional residual capacity by 11.1% (−21.6%; 1.2%), and lung clearance index by 18.3% (−44.0%; -0.3%). Additionally, within-visit variability was substantially reduced with the corrected signals.

**Conclusion:** Inadequate signal-correction of the measured CO_2_ concentration in the EasyOne Pro LAB device leads to a non-systematic error in expired nitrogen concentrations and overestimation of test outcomes. Two-point calibration of the CO_2_ sensor may maintain accurate measurement of gas concentrations and overcome this error.

## Introduction

Over the past decade, the lung clearance index (LCI) from the multiple-breath washout (MBW) test has become an established endpoint for clinical surveillance and interventional trials in patients with cystic fibrosis (CF)^1-6^. During a multiple-breath nitrogen washout, subjects inhale pure oxygen (100% O_2_) to gradually wash out resident nitrogen (N_2_) from the lung. The expired volume required to reach 2.5% of the starting N_2_ concentration provides a sensitive measure of ventilation inhomogeneity (LCI) expressed in multiples of the functional residual capacity (FRC)^7^.

Currently, two commercially available N_2_-MBW devices are mainly used in clinical trials, the EasyOne Pro LAB (EasyOne; ndd Medzintechnik AG; clinicaltrials.gov identifier: NCT04138589, NCT03691779) and the Exhalyzer D (Eco Medics AG; clinicaltrials.gov identifier: NCT04537793)^8,9^. Both devices derive the nitrogen concentration indirectly based on molar mass and gas measurements, but there is poor agreement in outcomes between setups and the devices can not be used interchangeably^10-12^. Previous studies have suggested several factors that might contribute to the disagreement between devices, including i) algorithms used to compute outcomes^10,11^, ii) dead space volumes^11,12^, iii) impact of on-demand oxygen supply on breathing pattern^10,12^, and iv) potential sensor errors^12^.

We recently characterized a substantial sensor-crosstalk error in the Exhalyzer D device that causes an overestimation of expired nitrogen concentrations. Correction for this error substantially reduces the main outcomes FRC and LCI by 8.9% and 11.9%, respectively, in children^13,14^ and by 10.5% and 16.1%, respectively, in infants and toddlers^15^. Unlike the Exhalyzer D, which requires daily calibration of the CO_2_ and O_2_ sensors^16^, the EasyOne device features a CO_2_ sensor advertised to provide accurate measurements without regular calibration^17^. Rather than being scaled from a two-point gas calibration, the EasyOne device assumes a fixed respiratory quotient (RQ) between patients and throughout the washout, and scales the measured CO_2_ signal to meet a set RQ condition using a correction factor (CO_2_-gain factor). The CO_2_-gain factor is used to correct the measured carbon dioxide signal for potential long-term changes in sensor output.

Due to the natural variability of the RQ, we hypothesized that correcting the measured CO_2_ signal based on an assumed and fixed RQ will result in inaccurate CO_2_ and N_2_ concentrations in the majority of measurements. In this study we investigated the accuracy of the RQ-based adjustment of the EasyOne CO_2_ sensor and assessed the impact of a corrected and an uncorrected CO_2_ concentration on MBW outcomes in healthy children and adults and children with lung disease.

## Methods

### Study Design and population

This was an experimental study to assess the accuracy of the measured and corrected CO_2_ concentration and its impact on MBW outcomes in the EasyOne device (ndd Medizintechnik AG, Zurich, Switzerland). We included MBW data from healthy children (Basel-Bern Infant Lung Development (BILD) cohort^18^), healthy adults, and children and adolescents diagnosed with CF or primary ciliary dyskinesia (PCD) lung disease (Table 1). The Ethics Committee of the Canton of Bern, Switzerland approved the study protocol (2019-01072, 2019-01591) and participants or parents gave written consent.

**Table 1.** Study population characteristics. Displayed as mean (SD) if not indicated otherwise. Age on test date as mean (min; max). Abbreviations: CF: cystic fibrosis; PCD: primary ciliary diskinisia; LCI: lung clearance index [turnover].

|  | Healthy controls |  | Lung disease |
| --- | --- | --- | --- |
|  | Children | Adults | Children |
| **Subjects** (female) [n] | 8 (6) | 10 (5) | 8 (5) |
| CF [n] | - | - | 7 |
| PCD [n] | - | - | 1 |
| **Age** [years] | 11.0 (8.6; 13.9) | 32.4 (27.1; 41.7) | 14.4 (11.0; 16.9) |
| **Weight** [kg] | 37.1 (5.8) | 71.9 (16.0) | 51.3 (8.7) |
| **Length** [cm] | 1.47 (0.1) | 1.76 (0.1) | 1.62 (0.1) |
| **LCI** [TO] | 7.37 (1.25) | 6.74 (0.8) | 8.56 (1.6) |

### Measurement principle and sensor characteristics

Applying Dalton’s law of partial pressures, the EasyOne device (ndd Medizintechnik AG) computes an indirect nitrogen concentration based on the measurement of molar mass (MM) and carbon dioxide (CO_2_)^19^. Under the assumptions that i) only the four gases nitrogen, argon, carbon dioxide, and oxygen vary in concentration during the washout and ii) that nitrogen and argon are always present in an assumed fixed ratio, a nitrogen concentration can be computed by solving the equations 1 and 2^20^.

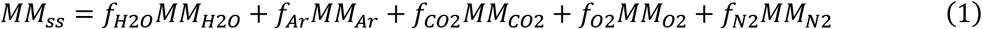

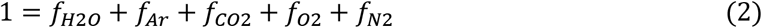

**Equations 1 and 2. Computation of nitrogen concentration based on measured molar mass and carbon dioxide**. Assuming that i) only the four gases N_2_, Ar, CO_2_ and O_2_ vary in concentration and ii) that N_2_ and Ar are always present in an assumed fixed ratio, the equations 1 and 2 are solved for the unknown concentrations of N_2_, Ar, and O_2_. Abbreviations: MM_ss_: sidestream molar mass, f: fraction, MM: molar mass, H_2_O: water vapour, Ar: argon, CO_2_: carbon dioxide, O_2_: oxygen, N_2_: nitrogen.

The sensors for MM and CO_2_ require a calibration to provide a stable and accurate signal over time^20^. For the MM sensor, a two-point calibration is performed during each measurement, using the known molar mass values of pure oxygen and room air, which are encountered during and after an automatic oxygen pulse before each measurement. For the CO_2_ sensor, such a calibration is not possible due to a lack of two distinct points of known CO_2_ concentration. Instead of performing a calibration for the CO_2_ sensor, the EasyOne device calculates a correction factor for the CO_2_ signal (CO_2_-gain factor) based on the assumption that the respiratory quotient (equation 3) during the first breaths before the wash-out start (pre-phase) is equal to a fixed value across the measurement and between patients. The CO_2_ signal in each measurement is then adjusted to meet this condition for the RQ^20^.

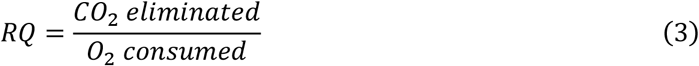

**Equation 3. Respiratory quotient**. Abbreviations: RQ: respiratory quotient, CO_2_: carbon dioxide, O_2_: oxygen.

### Validation of CO_2_ concentration

To validate the RQ-based rescaling of the measured CO_2_ signal, we collected standard N_2_-MBW measurements using the EasyOne Pro LAB (ndd Medizintechnik AG, Zurich, Switzerland) as recommended by the manufacturer^20^ and current consensus guidelines^21^ and added a pulse of a technical gas mixture (reference) at the end of each measurement (Figure 1). We compared the CO_2_ concentration during the gas pulse reported by WBreath analysis software (v3.54.0, ndd Medizintechnik AG, Zurich, Switzerland) with the known concentration of the reference gas mixture. The technical gas mixture contained 5% CO_2_, 95% O_2_, and no N_2_ with a precision of ±1% (Carbagas AG, Muri bei Bern, Switzerland). We then recalculated the CO_2_-gain factor required to set the measured CO_2_ concentration to the reference gas concentration, and re-analyzed the gathered raw data with corrected CO_2_-gain factors to determine the impact on CO_2_ and N_2_ concentrations and MBW outcomes (calibrated CO_2_). In addition, we performed a re-analysis without adjustment of the measured CO_2_ signal (fixed CO_2_-gain factor of 1.0).

**Figure 1.**
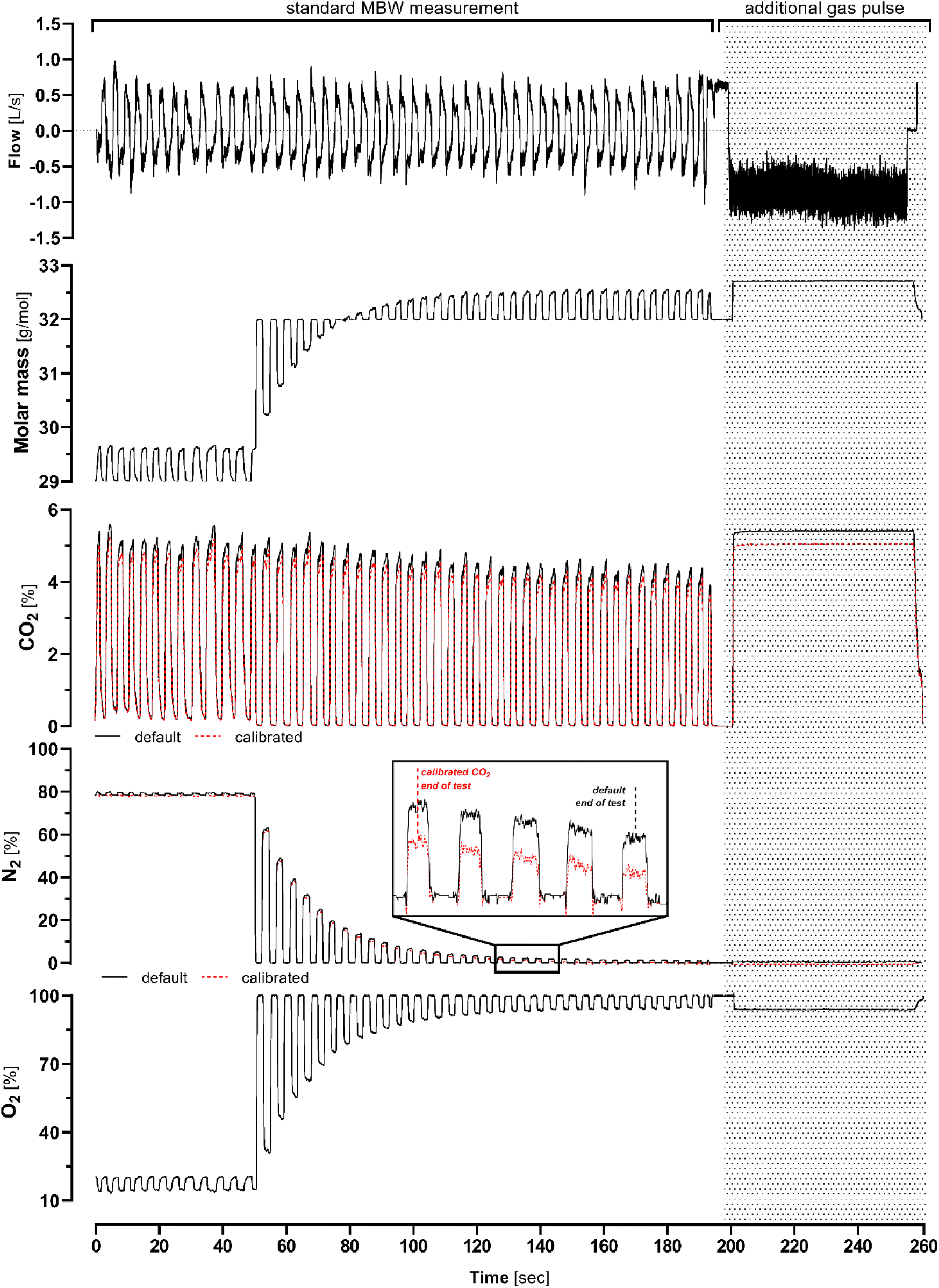
Signal traces of a MBW trial with reference gas pulse. Displayed are flow [L/s], sidestream molar mass [g/mol], default (black solid) and calibrated (red dashed) O_2_-corrected CO_2_ [%], default and calibrated N_2_ [%], and O_2_ [%] signals including a gas pulse with a technical gas mixture (shaded area) of a healthy adult. Default signals were analyzed as recommended while calibrated CO_2_-signals were recalculated based on the known CO_2_-concentration of the reference gas with adjusted CO_2_-gain values for CO_2_ (WBreath v3.54.0). In the N_2_ panel, end of test marks the first breath of the washout with an end-expiratory tracer gas-concentration below the end-of-test criterion of 1/40^th^ of the initial tracer gas concentration (normalized 2.5%) for default and CO_2_-calibrated signals, respectively. Reanalysis with calibrated CO_2_ values resulted in a decrease in nitrogen concentration and an earlier end-of-test (e.g. reaching 1/40^th^ of the initial tracer gas concentration five breaths earlier (calibrated CO2 end-of-test) compared to the default end-of-test.

### Data Analysis

To assess the accuracy of the RQ-based adjustment of the CO_2_ sensor we used the EasyOne CO_2_ sensor to measure the CO_2_ concentration of a high precision technical gas mixture. We calculated the mean CO_2_ concentration of the sensor reading over a 30-second pulse while systematically excluding any artifacts at the start/end of the pulse (e.g. sensor rise times). We assessed the impact of a measured and corrected CO_2_ signal on MBW outcomes (LCI, FRC, CEV) by comparing the outcomes of a default analysis (default; WBreath 3.54.0, ndd Medizintechnik AG, Zurich, Switerland) with outcomes of a corrected analysis (with modified CO_2_-gain factors; calibrated CO_2_) as well as without correction of the CO_2_ signal (fixed CO_2_-gain) using paired t-tests and Bland-Altman plots. Intergroup differences were compared using unpaired t-tests. Within-visit repeatability (standard deviation) was compared using a variance ratio test. Statistical analysis was performed using Stata 16.1 (StataCorp, College Station, USA) and GraphPad Prism 8 (GraphPad Software, San Diego, USA). A p≤0.05 was considered significant.

## Results

We assessed the CO_2_ concentration in 72 trials from 26 study visits in healthy children, healthy adults, and children with lung disease (Table 1). Four trials (three in healthy children, one in children with lung disease) were excluded from this analysis (three due to irregular breathing; one due to a possible leak).

### Error in CO_2_ concentration and resulting N_2_ concentration

We found that the RQ-based adjustment of the measured CO_2_ concentration used in the EasyOne device results in a random error of the CO_2_ concentration (range: -0.1 to 0.8%; Figure 1 and 2A). Based on known concentrations during the reference gas pulse (5% CO_2_, 95% O_2_, 0% N_2_), we derived corrected CO_2_-gain factors and re-analyzed the gathered raw data. Due to the indirect measurement principle, the error in CO_2_ concentration led to a non-systematic error in nitrogen concentration (Figure 2B). In 64/72 (88.9%) measurements, the nitrogen concentration was overestimated (range: -0.7 to 1.7%).

**Figure 2.**
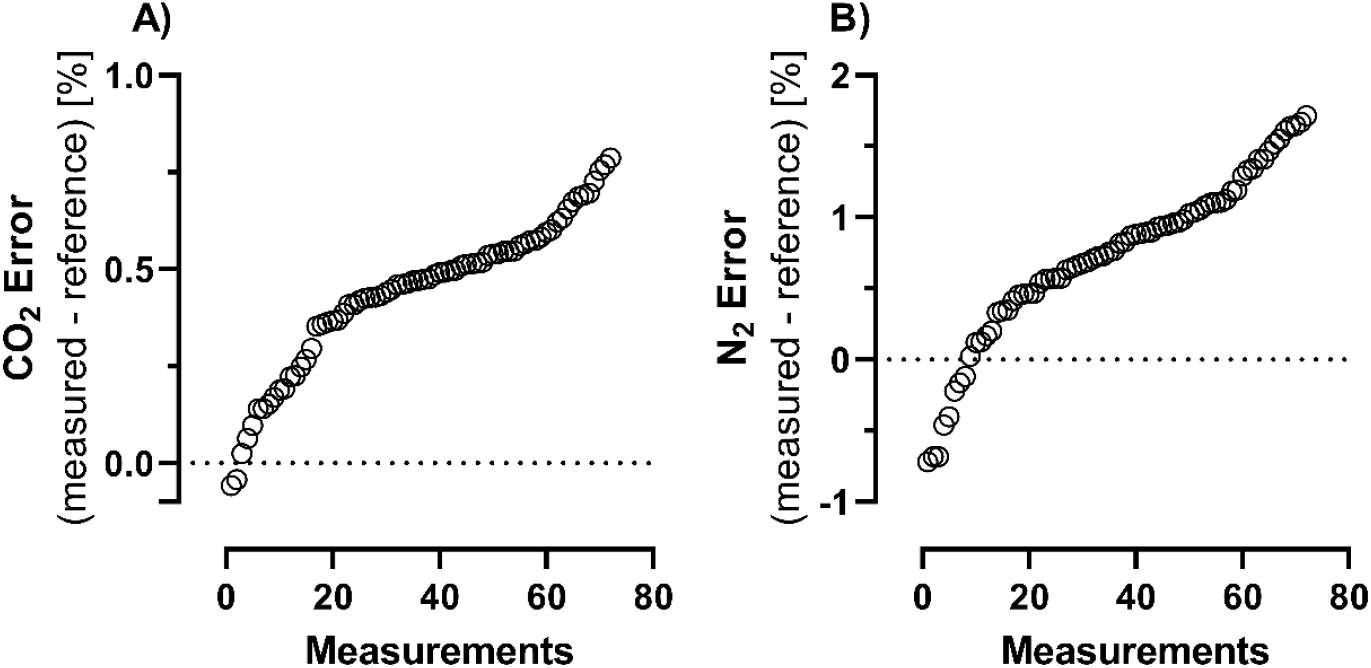
CO_2_ and N_2_ error. Shown are the absolute differences between the measured and known reference gas concentrations during the gas pulse for CO_2_ (A) and derived N_2_ (B) for 72 measurements.

### Impact of correct CO_2_-concentrations on MBW outcomes

Application of corrected CO_2_-gain factors had a significant impact on all MBW outcomes (calibrated CO_2_; Figure 3 and table 2). Following the correct CO_2_-calibration, the mean (range) cumulative expired volume (CEV) decreased by 27.1% (−58.1% to 1.0%), functional residual capacity (FRC) decreased by 11.1% (−21.6% to 1.2%), and LCI decreased by 18.3% (−44.0% to -0.3%). Overall, the change in outcomes following calibration of the CO_2_ correction was dependent on the magnitude of the outcomes themselves for both LCI and FRC (Figure 4). For most measurements, the lower corrected end-expiratory nitrogen concentration resulted in an earlier end of test (i.e., reaching 1/40^th^ of the initial N_2_ concentration) and thus a lower CEV. Lower N_2_ concentrations throughout the measurement caused a decrease in FRC. Re-analysis without correction of the measured CO_2_ signal (no CO_2_ correction; using a fixed CO_2_-gain factor of 1.0) resulted in small, yet statistically significant differences to the calibrated CO_2_ correction: mean CEV differed by 3.2%, mean FRC by 1.2%, and mean LCI by 1.9% (Figure 3 and supplemental table 1).

**Table 2.** Impact of calibrated CO_2_ correction on MBW outcomes. Visit means of LCIao, FRC, and CEV as reported in WBreath analysis software (v3.54.0) with default and calibrated (recalibrated to the reference CO_2_-concentration. Statistics: paired t test. Abbreviations: LCIao: lung clearance index [turnover] at airway opening; FRC: functional residual capacity [ml/kg]; CEV: cumulative expired volume [l].

|  |  |  | Default | Calibrated | Difference |  |  |  |  |  |
| --- | --- | --- | --- | --- | --- | --- | --- | --- | --- | --- |
|  |  | n | mean | mean | mean [TO] | range [TO] | mean [%] | range [%] |  | p value |
| **LCI [TO]** | HC adults | 10 | 6.74 | 5.81 | -0.93 | -1.8; 0.0 | -13.81 | -22.50; -0.25 |  | **0.0010** |
|  | HC children | 8 | 7.21 | 6.03 | -1.18 | -3.0; -0.5 | -16.41 | -35.55; -7.68 |  | **0.0043** |
|  | LD children | 8 | 8.56 | 6.09 | -2.47 | -4.5; -0.8 | -28.87 | -43.98; -10.28 |  | **0.0013** |
| **FRC [mL/kg]** | HC adults | 10 | 44.90 | 41.43 | -3.47 | -8.5; 0.5 | -7.73 | -14.77; 1.23 |  | **0.0015** |
|  | HC children | 8 | 39.22 | 34.91 | -4.31 | -5.7; -2.0 | -10.99 | -17.85; -4.29 |  | **0.0000** |
|  | LD children | 8 | 38.76 | 32.88 | -5.88 | -10.4; -2.3 | -15.17 | -21.58; -6.07 |  | **0.0004** |
| **CEV [L]** | HC adults | 10 | 24.04 | 18.84 | -5.20 | -15.5; 0.2 | -21.63 | -33.93; 0.96 |  | **0.0059** |
|  | HC children | 8 | 11.49 | 8.68 | -2.81 | -5.0; -1.9 | -24.45 | -48.05; -11.78 |  | **0.0001** |
|  | LD children | 8 | 19.35 | 11.48 | -7.87 | -15.8; -2.8 | -40.69 | -58.06; -16.23 |  | **0.0026** |

**Figure 3.**
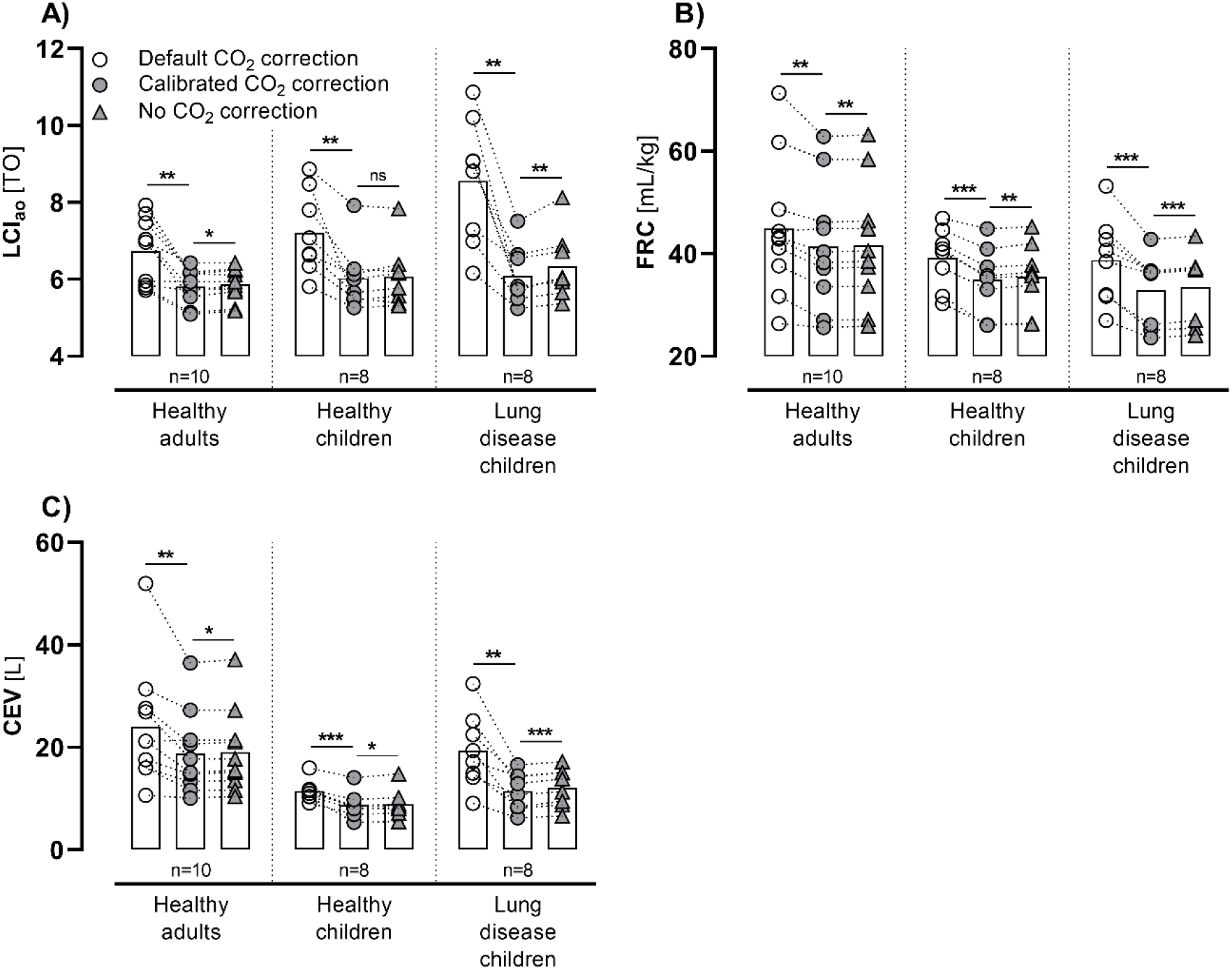
Impact of a corrected and uncorrected CO_2_ concentration on main MBW outcomes. Visit means of LCIao, FRC, and CEV as reported in WBreath analysis software (v3.54.0) with default CO_2_ correction (hollow circles), calibrated CO_2_ correction (recalibrated to the reference CO_2_ concentration (grey circles)), and measured CO_2_ without correction and a fixed CO_2_-gain factor of 1.0 (no CO_2_ correction (grey triangles)). Statistics: before-after plots with mean; paired t test: *p≤0.05, **p≤0.01, ***p≤0.001. Abbreviations: LCI_ao_: lung clearance index at airway opening [turnover]; FRC: functional residual capacity [ml/kg]; CEV: cumulative expired volume [l]; ns: no significant difference.

**Figure 4.**
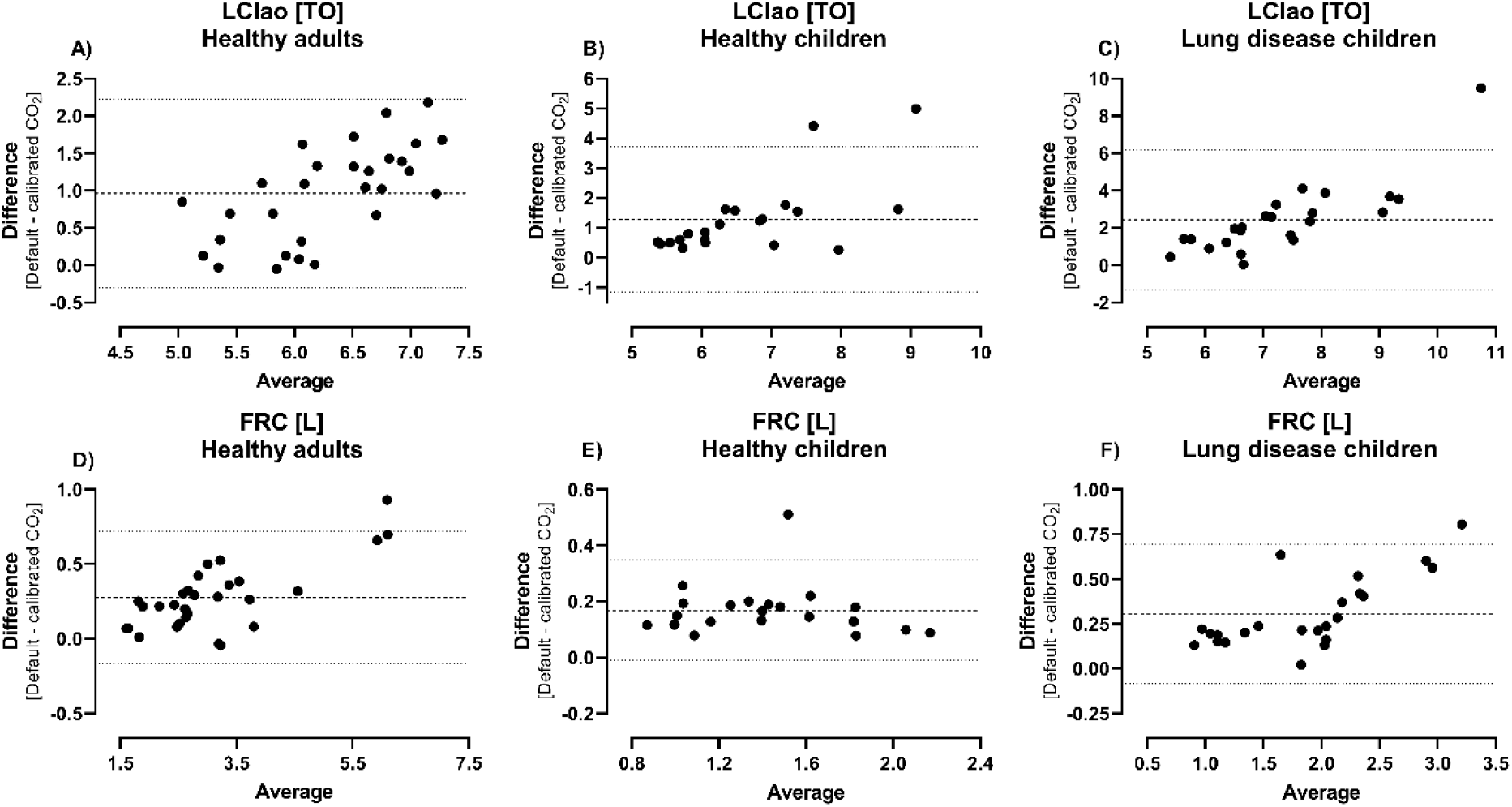
Bland-Altman plots of LCI (A-C) and FRC (D-F) agreement between default and calibrated CO_2_ concentration. The bold dashed line represents the bias, the fine dashed lines the upper and lower limits of agreement (mean difference ± 1.96 SD).

Across all study groups, within-visit variability was substantially reduced when using the calibrated CO_2_ concentration (SD_default_=1.7, SD_calibrated CO2_=0.7, p≤0.001; Figures 5A and B). Likewise, the uncorrected CO_2_ concentration (fixed CO_2_-gain) resulted in a smaller intra-test variability compared to the default CO_2_ concentration (SD_default_=1.7, SD_uncorrected CO2_=0.8, p≤0.001; Figures 5A and C).

**Figure 5.**
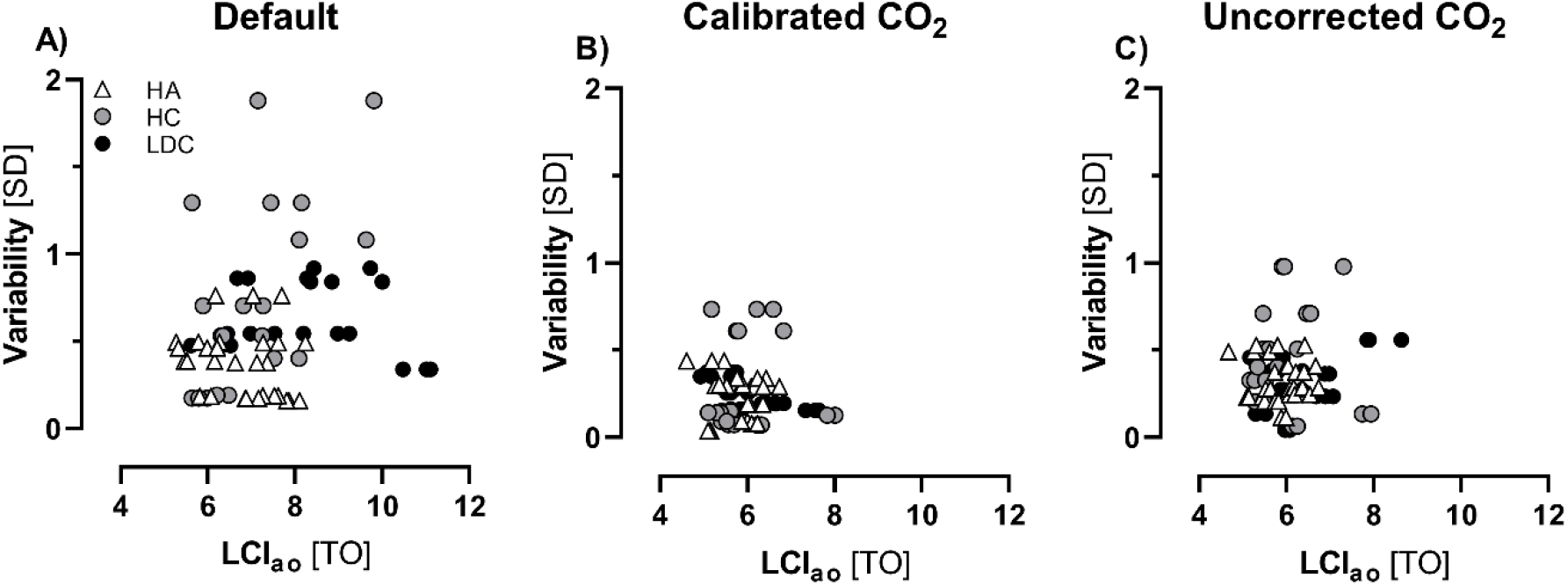
Change in within-visit variability. Within-visit variability (SD) was substantially reduced when analyzed with calibrated CO_2_ concentrations (recalibrated to the reference CO_2_-concentration (B, calibrated CO_2_)), as well as uncorrected CO_2_ concentrations and a fixed CO_2_-gain factor of 1.0 (C, uncorrected CO_2_) compared to the recommended analysis (A, default). Abbreviations: HA: healthy adults (n=10); HC: healthy children (n=8); LDC: lung disease children (n=8); LCI_ao_: lung clearance index at airway opening [turnover].

## Discussion

The objective of this study was to investigate the accuracy of the RQ-based adjustment of the measured CO_2_ concentration and the impact of corrected CO_2_ concentration on MBW outcomes in healthy children and adults and children with lung disease. We report a significant, non-systematic error in the EasyOne Pro LAB MBW device which in most measurements resulted in an overestimation of expired nitrogen concentrations, and consequently MBW results. Additionally, this error resulted in high within-visit variability in outcomes, especially in children.

### Sensor accuracy

We characterized the RQ-based signal correction and resulting non-systematic error in carbon dioxide and nitrogen concentration. Previous validation studies using *in-vitro* lung models did not specifically examine the influence of continuous and/or physiological CO_2_ concentrations over the course of the washout^10,12^, mainly because of two reasons: i) additional CO_2_ causes a volume drift when added to a Plexiglas model, and ii) a starting concentration of (e.g. 5%) CO_2_ in the lung compartment of a lung model without continuously added CO_2_ influx will washout over the first few breaths of the washout, thus missing the vulnerable period at the end of the washout where small errors may have a big impact on derived nitrogen concentrations. This made it obviously difficult to identify the signal-correction error of the CO_2_ sensor as there was no reference CO_2_ concentration for validation.

The observed error in our EasyOne device resulted in N_2_ concentrations between -0.7 to +1.7% during a gas mixture pulse where no nitrogen was present, thus exceeding the recommended measurement accuracy outlined by the American Thoracic Society (ATS) and the European Respiratory Society (ERS) of relative 5% accuracy at the test end^21^. Furthermore, the observed error in CO_2_ concentration (range: -0.1 to 0.8%) during the gas mixture pulse exceeded sensor accuracy of 0.15% specified by the manufacturer^17,22^.

Together with the reports of a substantial cross-sensitivity error in the Exhalyzer D MBW device^13^, these findings once more emphasize the need for more robust methods of validation of MBW devices. Ideally, an O_2_-consuming and CO_2_-producing *in-vitro* lung model should resemble physiological conditions for a wide range of lung volumes while allowing direct comparison of outcomes. Certainly, MBW devices based on an indirect measurement principle (e.g. Exhalyzer D, Eco Medics AG, and EasyOne Pro LAB, ndd Medzintechnik AG) should be transparent to the operator and enable the validation of measured gas concentrations which in turn are used to derive tracer gas concentrations.

### Correction of signal correction error

The respiratory quotient-based signal correction error observed in this study seems to be non-systematic. While the application of a known reference gas mixture to calibrate the measured CO_2_ concentration is an option for future measurements, it is not possible to correct for this error in the retrospective analysis of existing data. Due to the non-systematic nature of the error and the unknown “true” RQ of each trial, there is no correction function which could be applied to existing datasets at the moment. However, in our device, not applying such an adjustment at all and instead using a fixed CO_2_-gain factor substantially reduced the variability and resulted in smaller differences compared to the calibrated CO_2_ correction. Thus, a center or device-specific fixed CO_2_-gain factor could potentially be determined and used for retrospective corrections to existing datasets within a certain margin of error. However, this does not account for unknown long-term changes in sensor output over the history of the device.

### Effect size of correct calibration

The reference gas pulse at the end of each measurement made it possible to correct the CO_2_ signal to a known concentration by applying individual CO_2_-gain factors. As the nitrogen concentration was overestimated in the majority of measurements, correction of the CO_2_ signal resulted in substantially reduced MBW outcomes overall with a large effect size ranging between –44.0% and -0.3% for LCI. The effect was more pronounced in children than in adults and even greater in children with lung disease, where the mean (range) LCI changed by -28.9% (−44.0%; -10.3%). It is unclear whether this was due to higher LCI values or related to different RQs in patients with lung disease.

Previous studies reported significant differences in main MBW outcomes between the commercial EasyOne and Exhalyzer D MBW devices and FRC from other lung function tests (e.g. body plethysmography)^10,12^. Differences in the underlying algorithms for signal processing of raw data and the calculation of MBW outcomes, as well as potential effects of on-demand oxygen supply via variable bias flows on the breathing pattern have been the focus of attention. However, these factors could not sufficiently explain consistently lower lung volumes in the EasyOne when compared to other lung function tests (e.g. body plethysmography) and the Exhalyzer D or, depending on the study, varying differences in LCI^11,12^. Given that the correction of the recently described cross-sensitivity error in the Exhalyzer D device resulted in a decrease of FRC by 8.9% and that the correct CO_2_ signal in the EasyOne device resulted in an 11.1% reduction of FRC, the corrected values of both devices could potentially narrow the previously described difference^10,12^.

We found that it was not possible to correct CO_2_ and N_2_ concentrations simultaneously. A summary on remaining differences in MBW outcomes when analysed for correct CO_2_ or N_2_ concentrations is provided in the online supplement. Possible causes for the remaining difference include i) an erroneous drift correction of the molar mass signal, ii) a water vapor dependence in the argon-to-nitrogen ratio, iii) an O_2_-CO_2_ crosstalk error, or iv) an error in any of the underlying constants. Therefore, additional work is needed to systematically investigate these algorithms.

### Impact on clinical interpretation and ongoing studies

Our findings call into question previously published results obtained with the EasyOne MBW device. The respiratory quotient-based signal correction error observed in this study was non-systematic and there is no validated correction function for existing datasets at the moment. While the use of a device-specific fixed CO_2_-gain factor may improve the range of the observed error, this retrospective correction might not be ideal for older set-ups with potential drifts in CO_2_ sensor output over time. Furthermore, the remaining difference in nitrogen concentration in measurements with corrected CO_2_ indicates yet another, currently unknown issue.

Based on our findings and personal communication with Dr. Christian Buess (CTO, ndd Medizintechnik AG, Zurich, Switzerland), further investigation of the remaining differences in gas concentrations could include measurements with multiple reference-gas pulses. In the meantime, however, an adapted calibration algorithm in an updated analysis software along with reference gas concentrations of i) 100% O_2_, ii) 95% O_2_ and 5% CO_2_, and iii) room air could provide the essential information to redetermine the molar mass constants relevant to the underlying signal corrections.

Importantly it is not known how much impact this non-systematic error might have on results of clinical trials, particularly if the RQ itself could be influenced by the intervention. Assuming e.g. that the RQ in patients with CF changes systematically upon novel therapies with triple combination modulators, it is theoretically possible that effect sizes assessed by LCI are systematically overestimated when measured using this device (clinicaltrials.gov identifier: NCT04138589, NCT03691779)^8^.

### Strengths and Limitations

Through detailed understanding of the underlying signal processing of the EasyOne device, we were able to characterize the impact of a signal-correction error of the CO_2_ sensor on the clinical outcomes LCI and FRC for children and adults. However, we only had access to a single device and reference gas mixtures with finite precision to test the sensors and thus cannot make statements on other devices. However, as potential errors associated with applying a fixed RQ from the pre-phase to adjust the measured CO_2_ signal would apply to all devices, we are convinced that our results need to be considered for all setups. Due to the non-systematic nature of the signal-correction error, we were not able to develop a correction algorithm to correct for previously collected data.

## Conclusion

Inadequate signal-correction of the measured CO_2_ concentration in the EasyOne Pro LAB device leads to a non-systematic error in expired nitrogen concentrations and overestimation of clinical outcomes. A two-point calibration of the CO_2_ sensor with a reference gas may maintain accurate measurement of gas concentrations over time and overcome this error.

## Data Availability

All data produced in the present study are available upon reasonable request to the authors.

## Acknowledgements

We thank all our study participants and their families for allowing their MBW data to be used for research. Dr. Christian Buess (ndd Medizintechnik AG, Zurich, Switzerland) gave detailled advice on the measurement principle and algorithms for the computation of MBW outcomes.

## Author contributions and funding

MO and FW conception and design of research; MO data collection; MO, FW and BE analyzed data; MO, FW, PL, and KR interpreted results; MO prepared figures; MO drafted the manuscript; MO, FW, PL and KR edited and revised the manuscript; MO, FW, BE, PL, and KR approved the final version of the manuscript.

This project was funded by the Swiss National Science Foundation Grant Nr. 182719 (P. Latzin) and 168173 (K. Ramsey).

