## Supplemental material for "A non-systematic signal-correction error in a commercial multiple-breath washout device significantly impacts outcomes in children and adults"

### Online supplement

Marc-Alexander Oestreich[1,2†] (https://orcid.org/0000-0001-9641-3691)

Florian Wyler[1†] (https://orcid.org/0000-0002-1232-1392)

Blaise Etter[1] (https://orcid.org/0000-0002-8324-5008)

Kathryn A. Ramsey, PhD[1] (https://orcid.org/0000-0003-4574-6917)

Philipp Latzin, MD, PhD[1] (https://orcid.org/0000-0002-5239-1571).

[†]Authors contributed equally to this work as first author.

[1]Division of Paediatric Respiratory Medicine and Allergology, Department of Paediatrics, Inselspital, Bern University Hospital, University of Bern, Switzerland.

[2]Graduate School for Health Sciences, University of Bern, Switzerland.

**Corresponding author**:

Philipp Latzin, MD, PhD

Inselspital

Bern University Hospital

Freiburgstrasse 15, CH-3010 Bern, Switzerland

Impact of corrected and uncorrected $CO_2$ concentrations on main MBW outcomes

While the application of a calibrated $CO_2$-correction had substantial effects on main MBW outcomes (Table 1 of the manuscript), re-analysis without any correction of the measured $CO_2$ concentration resulted in comparable outcomes (supplemental table 1). Without correction, the mean (SD) cumulative expired volume (CEV) decreased by 24.9% (13.8%), functional residual capacity (FRC) decreased by 10.0% (5.3%), and LCI decreased by 16.7% (10.5%).

| | | | **Default** | **No correction** | **Difference** | | | | |
|---|---|---|---|---|---|---|---|---|---|
| | | n | mean | mean | mean [TO] | range [TO] | mean [%] | range [%] | p value |
| **LCI** [TO] | HC adults | 10 | 6.74 | 5.86 | -0.87 | -1.8 ; 0.0 | -12.98 | -22.88 ; -0.06 | **0.0017** |
| | HC children | 8 | 7.21 | 6.08 | -1.13 | -2.9 ; -0.3 | -15.72 | -34.38 ; -3.77 | **0.0059** |
| | LD children | 8 | 8.56 | 6.35 | -2.22 | -4.2 ; -0.6 | -25.88 | -40.74 ; -7.72 | **0.0017** |
| **FRC** [mL/kg] | HC adults | 10 | 44.90 | 41.59 | -3.31 | -8.2 ; 0.5 | -7.38 | -14.34 ; 1.14 | **0.0017** |
| | HC children | 8 | 39.22 | 35.45 | -3.77 | -5.4 ; -1.6 | -9.61 | -16.94 ; -3.46 | **0.0001** |
| | LD children | 8 | 38.76 | 33.51 | -5.24 | -9.8 ; -1.7 | -13.53 | -20.35 ; -4.44 | **0.0008** |
| **CEV** [L] | HC adults | 10 | 24.04 | 19.09 | -4.96 | -14.8 ; 0.2 | -20.62 | -33.93 ; 0.96 | **0.0067** |
| | HC children | 8 | 11.49 | 8.91 | -2.58 | -4.8 ; -1.2 | -22.46 | -46.51 ; -7.38 | **0.0004** |
| | LD children | 8 | 19.35 | 12.15 | -7.20 | -15.2 ; -2.1 | -37.20 | -54.95 ; -12.28 | **0.0039** |

**Supplemental table 1. Impact of calibrated $CO_2$ correction (A) and no $CO_2$ correction (B) on MBW outcomes.** Visit means of LCIao, FRC, and CEV as reported in WBreath analysis software (v3.54.0) with default and calibrated (A; recalibrated to the reference $CO_2$-concentration) and default and uncorrected $CO_2$ correction (fixed $CO_2$-gain factor of 1.0). Statistics: paired t test. Abbreviations: $LCI_{ao}$: lung clearance index [turnover] at airway opening; FRC: functional residual capacity [ml/kg]; CEV: cumulative expired volume [l].

### Impact of $N_2$-calibration on MBW outcomes

It was not possible to correct $CO_2$ and $N_2$ concentrations simultaneously. All MBW outcomes differed substantially when analysed for correct $CO_2$ or $N_2$ concentrations (supplemantal table 2 and supplemental figure 1). Following the re-calibration for correct nitrogen concentrations, the mean (SD) cumulative expired volume (CEV) decreased by 14.6% (16.2%), functional residual capacity (FRC) decreased by 5.9% (6.0%), and LCIao decreased by 9.9% (11.9%).

| | | | **Calibrated $CO_2$** | **Calibrated $N_2$** | **Difference** | | | | |
|---|---|---|---|---|---|---|---|---|---|
| | | n | mean | mean | mean [TO] | range [TO] | mean [%] | range [%] | p value |
| **LCI** [TO] | HC adults | 10 | 5.81 | 6.17 | 0.36 | 0.1 ; 0.5 | 6.22 | 1.24 ; 8.75 | **0.0000** |
| | HC children | 8 | 6.03 | 6.44 | 0.41 | 0.3 ; 0.7 | 6.83 | 4.48 ; 12.00 | **0.0001** |
| | LD children | 8 | 6.09 | 6.76 | 0.67 | 0.4 ; 1.1 | 11.06 | 7.63 ; 14.46 | **0.0000** |
| **FRC** [mL/kg] | HC adults | 10 | 41.43 | 43.15 | 1.72 | 1.4 ; 2.7 | 4.15 | 2.77 ; 6.35 | **0.0000** |
| | HC children | 8 | 34.91 | 36.74 | 1.83 | 1.0 ; 2.8 | 5.24 | 2.69 ; 7.48 | **0.0001** |
| | LD children | 8 | 32.88 | 34.99 | 2.11 | 1.7 ; 2.7 | 6.43 | 5.20 ; 8.05 | **0.0000** |
| **CEV** [L] | HC adults | 10 | 18.84 | 20.81 | 1.96 | 1.1 ; 4.3 | 10.42 | 4.08 ; 14.40 | **0.0001** |
| | HC children | 8 | 8.68 | 9.73 | 1.05 | 0.7 ; 1.4 | 12.06 | 8.15 ; 16.97 | **0.0000** |
| | LD children | 8 | 11.48 | 13.55 | 2.07 | 1.2 ; 3.0 | 18.01 | 14.16 ; 24.34 | **0.0000** |

**Supplemental table 2. Difference in retrospective calibration for $CO_2$ and $N_2$ on MBW outcomes.** Abbreviations: LCI: lung clearance index [turnover]; FRC: functional residual capacity; CEV: cumulative expired volume.

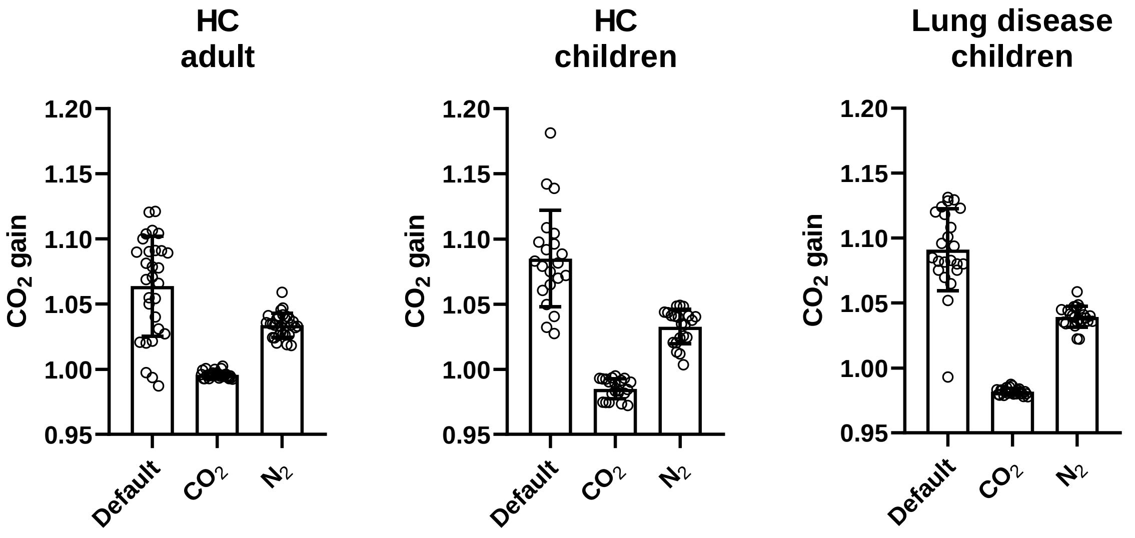


**Supplemental figure 1. Difference in $CO_2$-gain factor.** $CO_2$-gain factors of individual trials as reported in WBreath analysis software (v3.54.0) with recommended settings (default) and after calibration for the reference $CO_2$ and $N_2$ concentration. Statistics: mean with SD error bars; paired t test.

Double-pulse measurements

To investigate a possible drift in $CO_2$ concentration within a single measurement, we applied two reference gas pulses (5% $CO_2$, 95% $O_2$, 0% $N_2$) to an exemplary measurement of a healthy adult (supplemental figure 2) and compared measured $CO_2$ concentrations after a default analysis using WBreath analysis software (v3.54.0, ndd Medizintechnik AG, Zurich, Switzerland). The measured $CO_2$ concentrations remained stable before (5.52%) and after the washout (5.54%).

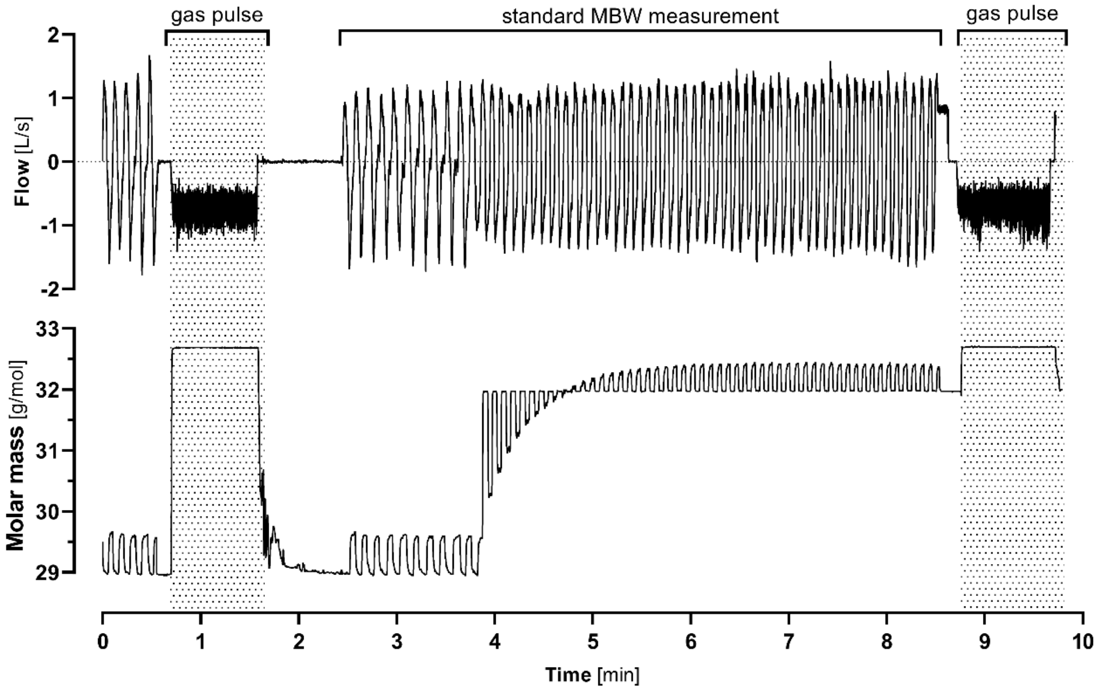


**Supplemental figure 2. Signal traces of a MBW trial with two reference gas pulses.** Displayed are flow [L/s] and sidestream molar mass [g/mol] including two gas pulse with a technical gas mixture (shaded area) of a healthy adult.
